# Candidate genes for IgA nephropathy in pediatric patients: exome-wide association study

**DOI:** 10.1101/2023.08.09.23293902

**Authors:** Anastasiia A. Buianova, Mariia V. Proskura, Valery V. Cheranev, Vera A. Belova, Anna O. Shmitko, Anna S. Pavlova, Iuliia A. Vasiliadis, Oleg N. Suchalko, Denis V. Rebrikov, Edita K. Petrosyan, Dmitriy O. Korostin

## Abstract

IgA nephropathy (IgAN) is an autoimmune disorder that commonly manifests during adolescence. This disease is believed to be a non-monogenic disorder related to variations in multiple genes involved in various biological pathways.

We performed the exome-wide association study of 70 children with IgAN confirmed by renal biopsy and 637 healthy donors to identify gene associations responsible for the disease. The HLA allele frequencies between the patients and healthy donors from the bone marrow registry of the Pirogov University were compared. We tested 78,020 gene markers for association, performed the functional enrichment analysis and the transcription factor binding preference detection.

We detected 333 genetic variants, employing three inheritance models. The most significant association with the disorder was observed for rs143409664 (*PRAG1*) in case of the additive and dominant models (P_BONF_ = 1.808 × 10^−15^ and P_BONF_ = 1.654 × 10^−15^, respectively) and for rs13028230 (*UBR3*) in case of the recessive model (P_BONF_ = 1.545 × 10^−9^). Enrichment analysis indicated the strongly overrepresented “immune system” and “kidney development” terms. The HLA-DQA1*01:01:01G allele (P = 0.0076; OR, 2.021 [95% CI, 1.322-3.048]) was significantly the most frequent among IgAN patients.

Here we characterized, for the first time, the genetic background of the Russian IgAN patients identifying the risk alleles typical of the population. The most prominent signals were detected in previously undescribed loci.

## Introduction

IgA nephropathy (IgAN) is one of the most common primary glomerulonephritides in the world both in adults and children [1]. Although it has been extensively studied for more than half a century, it still remains the leading cause of the end-stage kidney disease (ESKD) [2]. The clinical presentation of the disease is highly variable: from painless microhematuria to the rapidly progressing glomerulonephritis and ESKD [3–7]. From 20% to 40% of cases of IgAN progression to ESKD within 20 years from the onset require renal replacement therapy [8–11].

IgA nephropathy is diagnosed through kidney biopsy examination. It shows the deposition of IgA-containing immune complexes in the mesangium inducing mesangial cell proliferation and extracellular matrix accumulation [1]. At present, there are two basic views on the causes of the disease. Several groups suggest that IgAN is an autoimmune disease leading to the antibody-mediated destruction of the glomerular basement membrane. IgAN pathogenesis is complex and likely to involve several different pathways forming a complex network where infections may play a triggering role. Though in some cases, IgA nephropathy can precede the infection that induces a dysregulated immune response, IgA nephropathy itself is not an infectious disease [12].

Furthermore, in IgAN, the glycosylation of O-linked glycans in the hinge region of IgA1 is disrupted resulting in the high blood level of circulating galactose-deficient IgA1 (Gd-IgA1) and its abnormal clearance [13]. Gd-IgA1 has been shown to be highly heritable, therefore, the link between IgAN and Gd-IgA1 and its role in IgAN pathogenesis might provide novel insights into the pathogenic processes involved in IgAN [14]. Levy M et al. suggested a possible contribution of genetic predisposition to IgAN [15]. Their hypothesis is based on a certain geographical prevalence of the IgAN. Based on kidney biopsy, IgA nephropathy was diagnosed in 20% and 40% of children with glomerular diseases in Europe and Asia, respectively, whereas it was not frequently registered in the African population [7, 16].

The presence of genes involved in the immunity against intestinal pathogens is thought to be responsible for the high incidence of IgA nephropathy in Asia. Certain loci are associated with the risk of inflammatory bowel disease (IBD), the state of the intestinal epithelial barrier, and response to mucosal pathogens. The genetic predisposition strongly correlates with helminth diversity, suggesting a possible role for the host–intestinal pathogen interactions in IgAN geographic variations [17]. Familial cases of IgAN are also known and described supporting the genetic role in the disease etiology [18–22].

The role of gene-candidates in IgAN development was estimated by genome-wide association study (GWAS). In familial forms, a high risk of the disease was found to correlate with the following specific loci: *6q22–23 (IGAN1), 4q26–31 (IGAN2)* and *17q12–22 (IGAN3)* [23, 24]. In East Asia, the associated proteins contribute to the adaptive and innate immunity, IgA1 glycosylation, and the renin-angiotensin system [25], including haplotypes *HLA-DQ* and *HLA-DR*: HLA-DRB1*14:05:01 (belonging to DR*14), HLA-DRB1*03:01:01, HLA-DRB1*04, HLA-DQB1*03:01 [26–28]. Xia YF et al. showed the Megsin gene (*SERPINB7*) to be a major factor determining the predisposition to the disease and its progression in the Chinese population [29, 30]. Twenty-four candidate genes associated with IgAN were analyzed to identify their interactions. The cooperation between *C1GALT1-330G/T (*rs1008898) and *IL5RA31+197A/G* (rs340833) was found to be statistically significant (P < 0.001) for IgAN pathogenesis [31], whereas the combination of *P-selectin-2441a/G* with *CD14-159C/T* was associated with gross hematuria in IgAN patients. Moreover, the interaction of *TGF-b1 509T/C, P-selectin-2441A/G*, and *MCP-1 2518A/G* was found to influence the crescent formation [32]. The interaction between two key genes, *C1GALT1* and *ST6GALNAC2*, was also shown to affect the susceptibility to IgAN and disease progression [33]. In the UK population, a significant association with IgAN was detected on the chromosome 6 in the region of the MHC (P = 1 × 10^−9^) [34]. Five loci associated with susceptibility to IgAN were identified in Chinese patients: three distinct loci in the MHC region, a common deletion of *CFHR1* and *CFHR3 1q32* loci, and a locus *22q12 (HORMAD2)* [35]. In 2014, Kiryluk et al. revealed six haplotypes associated with a high risk of IgAN: already known variants in *ITGAM, ITGAX, VAV3* and *CARD9* genes, as well as two new haplotypes, *HLA-DQB1* and *DEFA* [17]. Ming Li et al. identified genes responsible for susceptibility to the disease and associated with IgAN, located in *17p13* and *8p23* [36], including the tumor necrosis factor (*TNFSF13*) and α-defensin (*DEFA*). rs660895 (*HLA-DRB1*) was found to correlate with the IgA serum level and proteinuria level [37]. The novel genes associated with IgAN included *ST6GAL1* at *3q27.3*, *ACCS* at *11p11.2*, and *ODF1-KLF10* at *8q22.3*. The *ITGAX-ITGAM (16p11.2*) association was confirmed being moderately replicated, and previously observed genes *VAV3(1p13)* and *CARD9 (9q34)* were detected as well.

Given that the genetic aspects of IgAN have previously demonstrated high heterogeneity in terms of the identified associations, we performed exome-wide association study (EWAS) employing our own clinical samples.

## Materials and Methods

### Ethics statement

All legal representatives as well as patients over 15 y.o. signed the appropriate informed voluntary consent to participate in the study. Local ethics committee of Pirogov Russian National Research Medical University approved this study on the 17^th^ of December, 2018 (Protocol No. 181).

### Patient cohort

Between 2019 and 2021, 70 children at the Russian Children’s Clinical Hospital had been observed at the nephrology department for at least 6 months. During laboratory tests, 4 ml of blood were sampled from the cubital vein into a test tube with EDTA for subsequent EWAS. In all patients, the diagnosis of primary IgA nephropathy was confirmed by kidney biopsy and further histological examination with immunofluorescence, electronic, and light microscopy.

### gDNA extraction

DNA isolation was performed using the DNeasy Blood and Tissue Kit (Qiagen) according to the manufacturer’s protocol. The extracted DNA was quantified with the Qubit dsDNA BR Assay system (Life Technologies), and its quality was assessed by 1% agarose gel electrophoresis.

### Library preparation and enrichment

DNA Libraries were prepared from 500 ng of genomic DNA using the MGIEasy Universal DNA Library Prep Set (MGI Tech) according to the manufacturer’s protocol. DNA fragmentation was performed by ultrasonication using Covaris S-220 with the average fragment length of 250 bp. Whole-exome enrichment of DNA library pools was performed according to a previously described protocol [38] using the SureSelect Human All Exon v7 probes (Agilent Technologies). The concentrations of DNA libraries were measured using Qubit Flex (Life Technologies) with the dsDNA HS Assay Kit following to the manufacturer’s protocol. The quality of the prepared libraries was assessed using Bioanalyzer 2100 with the High Sensitivity DNA kit (Agilent Technologies) according to the manufacturer’s instructions.

### Sequencing

The enriched library pools were further circularized and sequenced by a paired end sequencing using DNBSEQ-G400 with the DNBSEQ-G400RS High-throughput Sequencing Set PE100 following the manufacturer’s instructions (MGI Tech) with the average coverage of 100x Fastq files were generated using the basecallLite software from the manufacturer (MGI Tech).

### Control samples

We used exome sequencing data from 637 healthy Russian donors previously processed in our laboratory as a control dataset. To compare HLA allele frequencies, we used data on healthy donors from the Pirogov University register including 1849 individuals.

### Raw sequencing data analysis

The quality control of the obtained paired fastq files was performed by FastQC v0.11.9 [39]. Based on the quality metrics, fastq files were trimmed using BBDuk by BBMap v38.96 [40]. Reads were aligned to the indexed reference genome GRCh37 using bwa-mem2 v2.2.1 [41, 42]. SAM files were converted into bam files and sorted using SAMtools v1.9 to check the percentage of the aligned reads [43]. The duplicates in the obtained bam files were marked using Picard MarkDuplicates v2.22.4 [44] and excluded from further analysis.

We performed the quality control analysis on marked bam files with the following Agilent all-exon v7 target file “regions.bed”. For the samples that passed quality control (width of target coverage 10x≥95%), SNVs and indels were called using the bcftools mpileup software v1.9 [45], and vcf files were obtained for each sample. After variant calling, vcf files were normalized using vt normalize v0.5772-60f436c3 [46] and filtered based on the target regions expanded by +-100 base pairs flanking each end. Calling data were annotated using the InterVar software [47].

### Exome-wide association study

We analyzed DNA samples from 70 IgA nephropathy (IgAN) patients and 637 samples from healthy donors. Variants in vcf files (ver 4.2) were filtered based on coverage (threshold = 13) and quality (threshold = 20). InDel variants were normalized using vt (ver v0.5772-60f436c3) tool. All vcf files were merged by bcftools merge (ver 1.10.2). To obtain the list of genes containing sets of false positive variations, the genomic intervals were extracted from the gtf file of genome annotation (GRCh37.p13). Then, using bedtools (ver 2.27.1) [48], variations in these genomic intervals were excluded from the merged vcf files. Data were analyzed in the PLINK v1.90b6.24 software [49]. The deviation of sample heterozygosity levels was within 3 SD from the mean. The markers (both SNVs and indels) genotyped in less than 80% of the samples as well as individuals with less than 80% of genotyped markers were filtered out. In all samples, predicted sex coincided with the actual sex. Due to a small sample size, we chose a threshold of 0.0001 and a frequency of a minor allele of 0.1 for checking the Hardy-Weinberg equilibrium. After setting the threshold value PI_HAT = 0.02, we did not exclude samples from the analysis. Finally, 78,020 of 793,015 markers were subjected to further analysis. We created the additive, dominant, and recessive genetic models; p-value was corrected using the Bonferroni criterion (P_adj_ < 0.05).

### Functional enrichment analysis and transcription factor (TF) binding preference detection

The charts of gene enrichment analysis were created using the Metascape v3.5.20230101[50].

The eQTL analysis was performed using the following databases: Kidney eQTLs Atlas [51], NephQTL [52], and Database of Immune Cell eQTLs (DICE) project [53]. The cut-off for p-value was P < 0.05.

The STRING (https://string-db.org, date of access 12 April 2023) database was used to predict functional interactions of proteins. The search was restricted to “*Homo sapiens”* and the interaction score[>[0.4 were applied to construct the PPI networks.

We applied atSNP [54] to predict differential binding of a TF to a marker by labeling SNP-motif combinations with atSNP pval_diff < 0.05 as significant.

### HLA-typing

HLA typing was performed on the exome data with help of three programs: HLA-HD (ver 1.7.0) [55], HLAScan (ver 2.1.4) [56], Kourami (ver 0.9.6) [57] using the allele database IPD-IMGT/HLA (version 3.51.0) [58]. The obtained data on the alleles were transformed into the corresponding G-groups, and results were merged into the final version. In case different tools provided conflicting results, we chose the alleles identified using HLA-HD or Kourami and checked bam files in IGV [59].

### Statistical analysis

Principal component analysis (PCA) and allele frequency estimation were performed using PLINK v1.90b6.24 (Supplemental Figure 1). Manhattan and quantile-quantile (Q-Q) plot were generated with Python scripts. The one-way analysis of variance (ANOVA) test, Chi-squared test and Fisher’s exact test [60] were used to compare the differences between groups in RStudio 2022.02.0. The multiple comparison correction was conducted using the Bonferroni method: we multiplied p-values by the number of the alleles under consideration. Testing for the Hardy-Weinberg equilibrium was performed using the Arlequin 3.5 software [61]. All data were presented as mean ± standard deviation (SD) or median [Q1, Q3]. Data normality was checked using the Kolmogorov-Smirnov test.

## Results

### Clinical Characteristics

The clinical characteristics of 70 IgAN patients are summarized in table S1.

We found no significant effects of sex on the clinical features based on a one-way analysis of variance (ANOVA). The control sample had an even distribution of sexes, with 52.04% males and 47.96% female.

### Morphological analysis

Morphological data of kidney biopsy samples typical of their diagnosis are presented in Figure 1.

**Figure 1.**
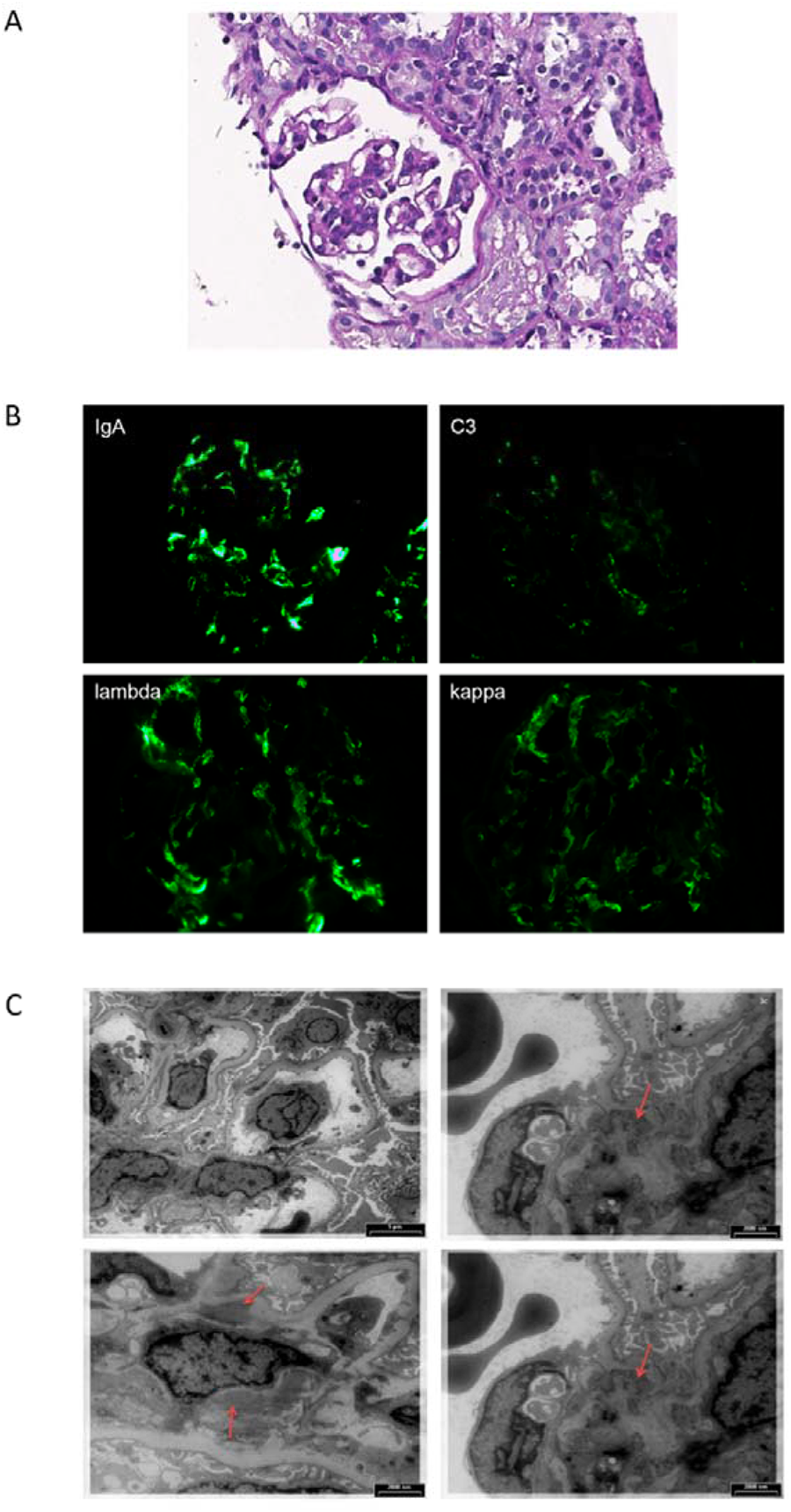
Renal biopsy of the typical patient with IgAN. Hematoxylin-eosin stain (magnification ×400) (A). Immunofluorescence microscopy, staining for IgA, C3, lambda, kappa (magnification ×400) (B). Electron microscopy. Red arrows indicate paramesangial deposits (C).

In the histological sample, matrix expansion and mesangial hypercellularity (up to 10 cells per a mesangial region) are presented (figure 1A). According to immunofluorescence: IgA, kappa, lambda in mesangium +++. IgG, IgM, C1q, fibrinogen – negative. С3 – ++ (figure 1B). The electron microscopy study reveals noticeable mesangial proliferation (4-6 nuclei) and massive multiple paramesangial deposits (figure 1C).

### Description of the sequencing data

The main quality control metrics for the whole-exome sequencing of IgAN patient samples are listed in table S2. Sex chromosome karyotype and results of analysis of the SRY gene coverage coincided with the data from the patient medical cards.

### Exome-wide association study of IgAN

IgAN patients were successfully matched with control subjects in case of the recessive model, as clearly shown by the small systematic deviation (λ = 1.196) of the observed distribution from the expected distribution under the null hypothesis which claimed the absence association in the Q-Q plot (figure 2A). The association p-value of multiple markers located on autosomes are shown on the Manhattan plots (figure 2B).

**Figure 2.**
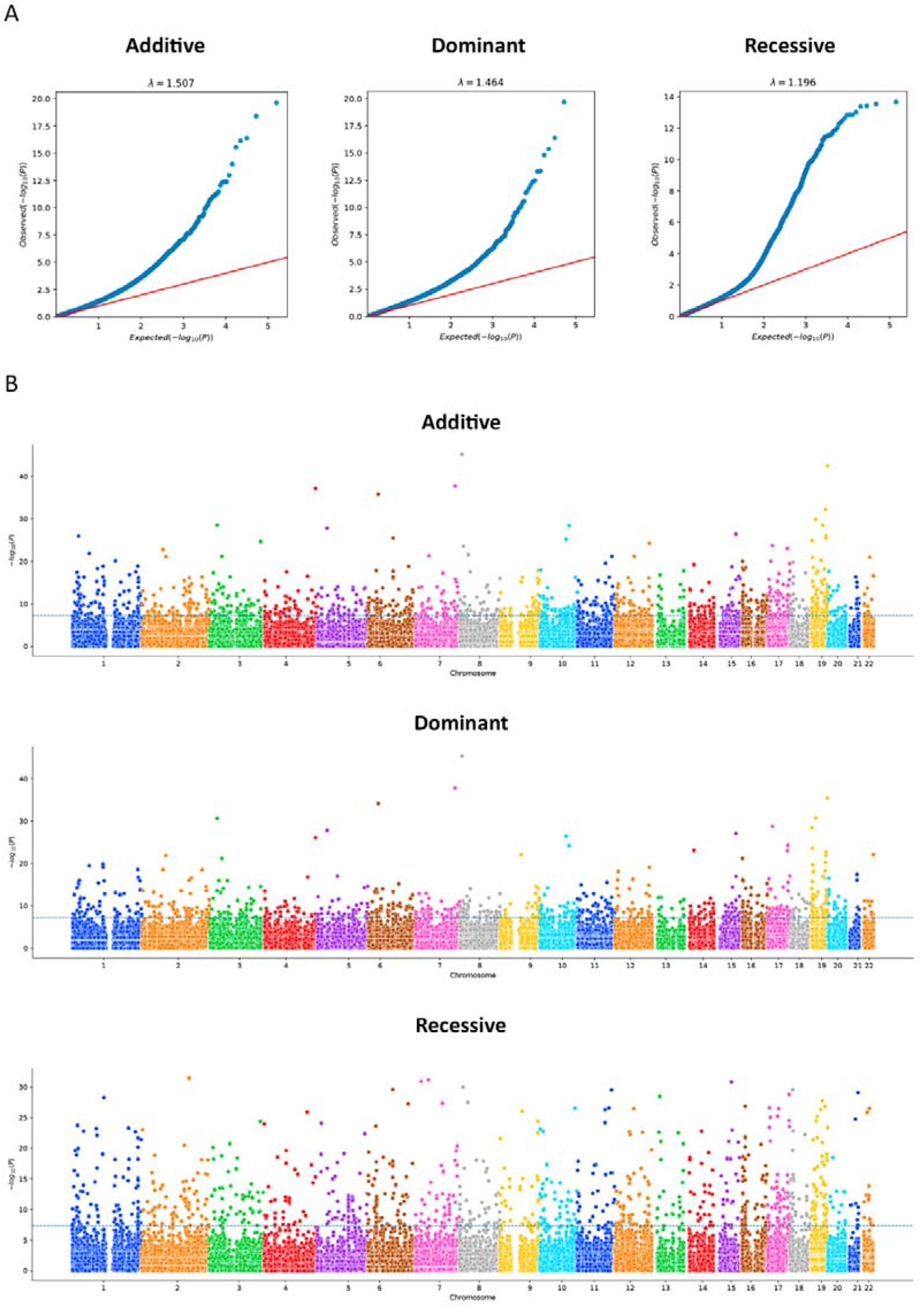
The Q-Q plots for association analysis. Each figure shows the expected (x-axis) and observed (y-axis) log (p-values) (A). The Manhattan plots for the exome-wide association study (EWAS) of IgA nephropathy (IgAN). The figure shows the p-value for the association with the disease (expressed as a negative logarithm of the p-value, y-axis) for each tested marker plotted against the chromosomal position of the markers (x-axis). The blue line at 6 represents the threshold for EWAS (−log_10_(1 × 10^−6^)) (B).

A total of 333 markers were identified after the p-value correction, including 52 insertions/ deletions. For each model, we showed the 10 most significant markers; with 24 of them not overlapping (table 1).

**Table 1.**
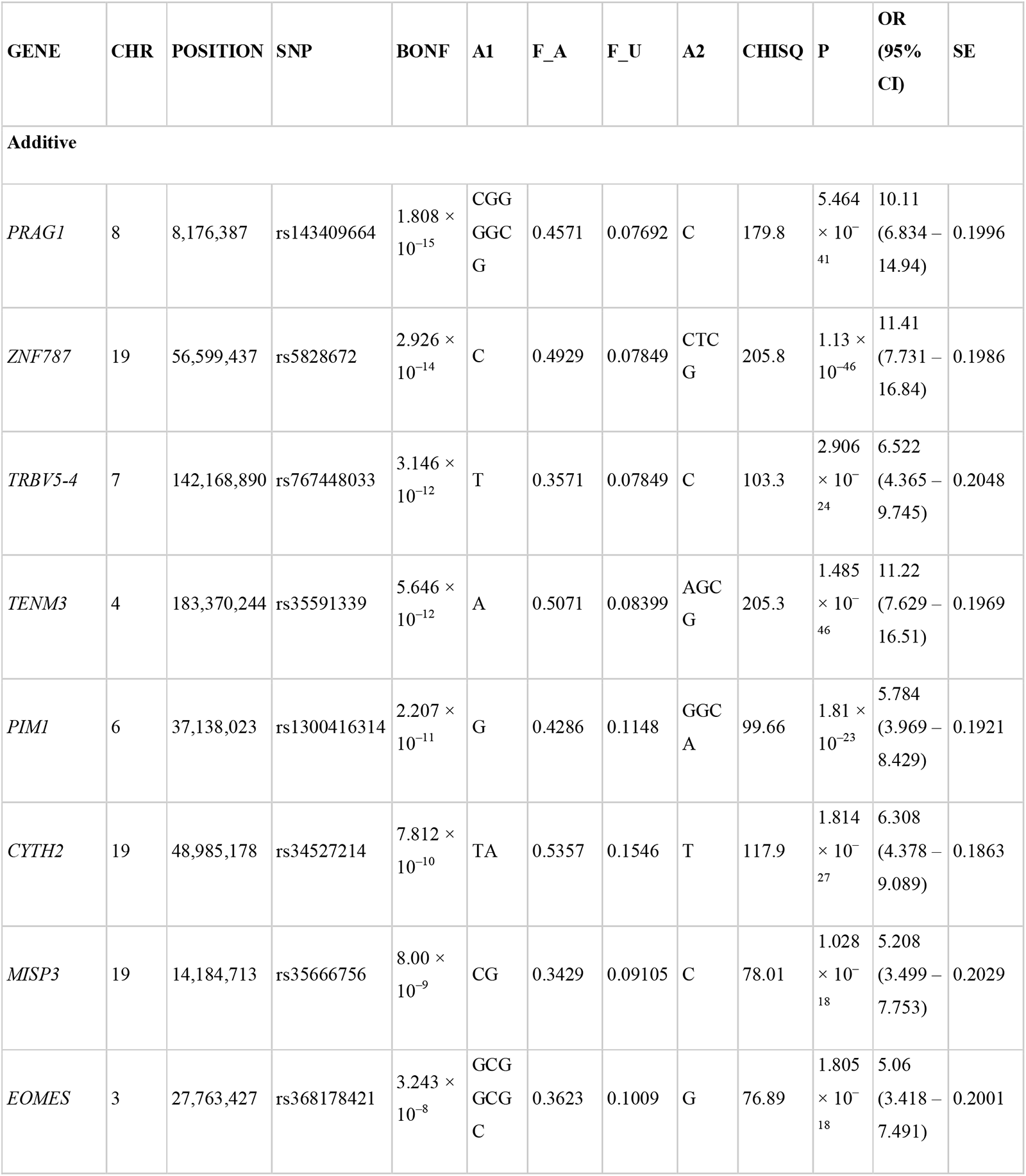
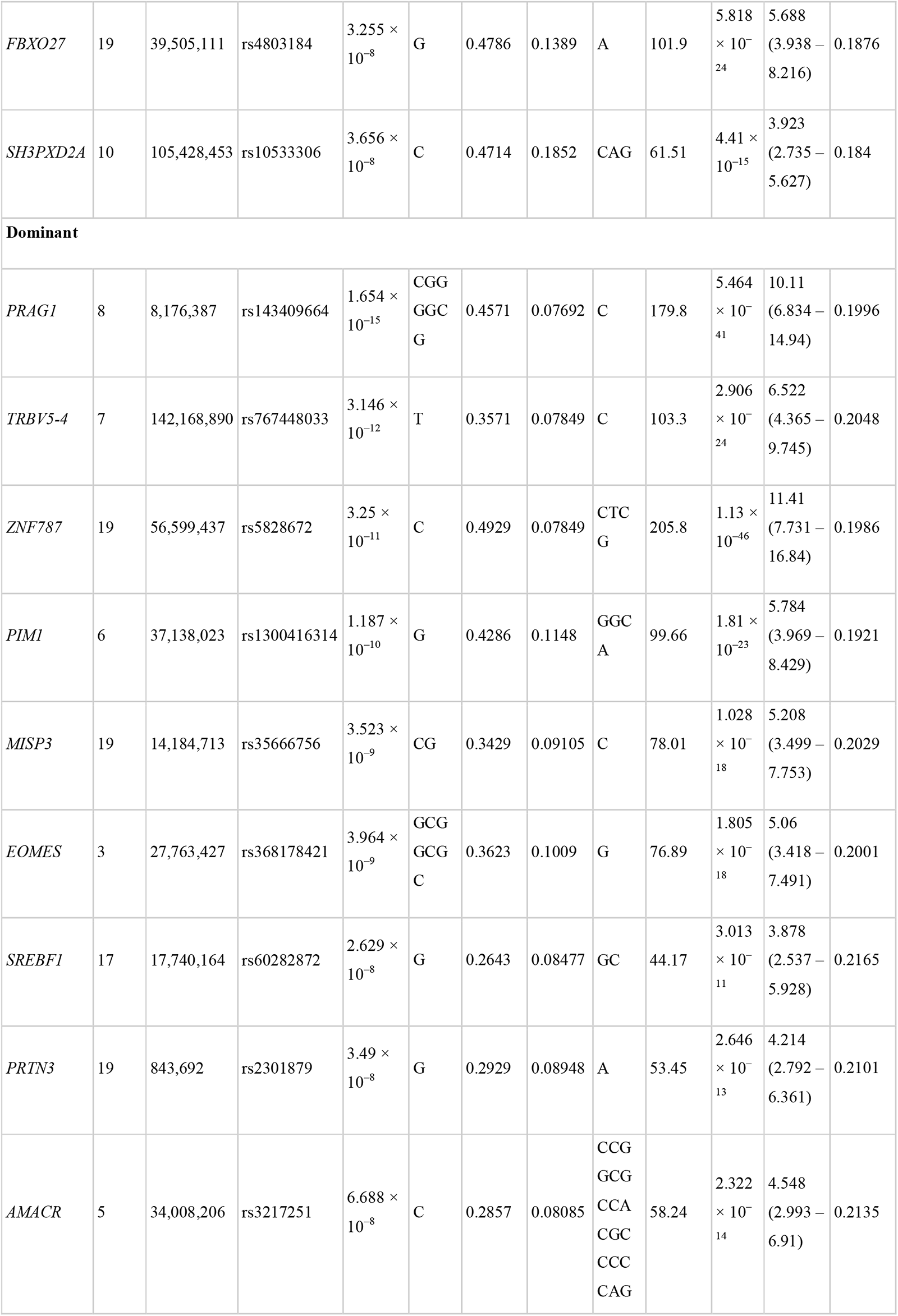
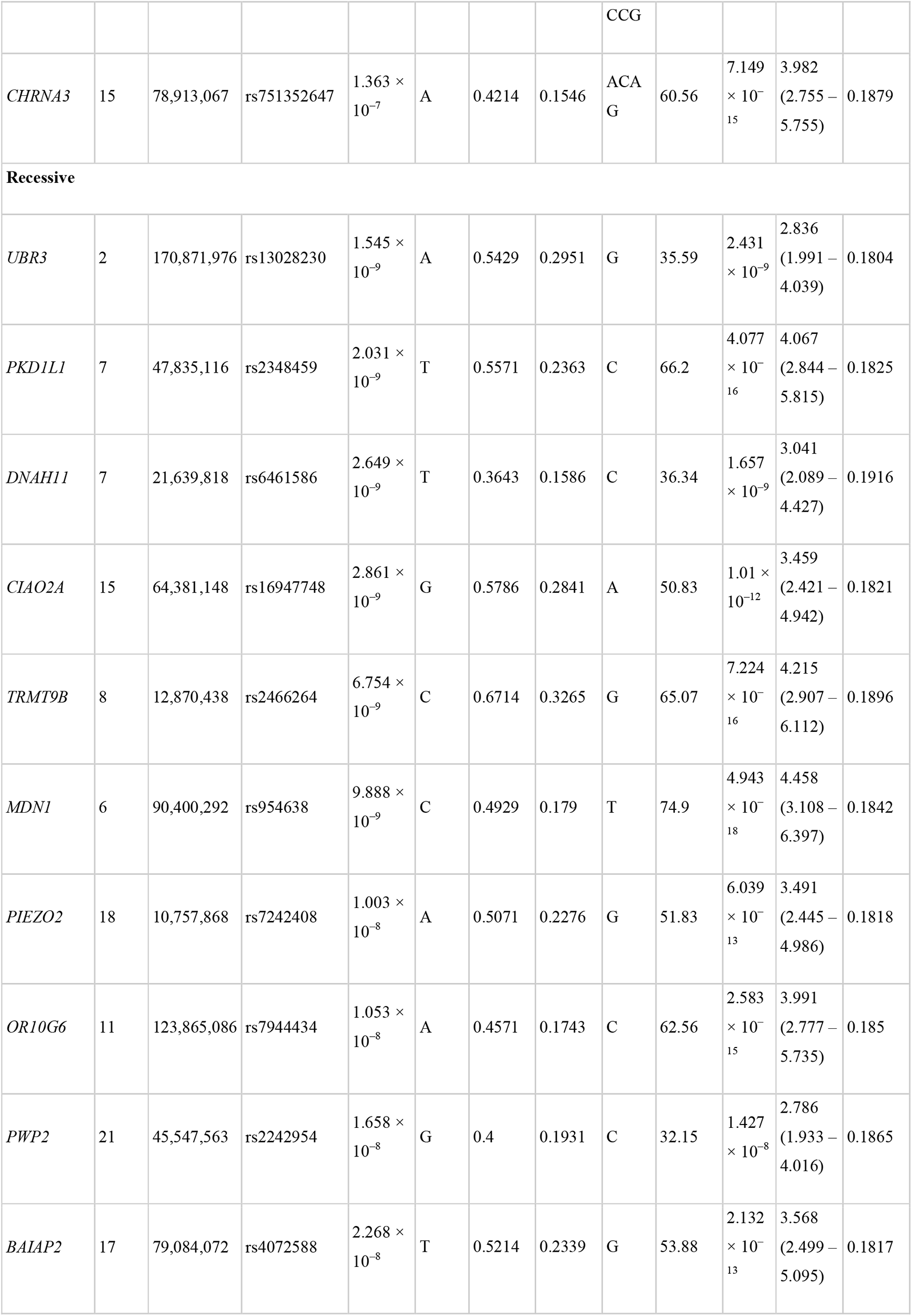
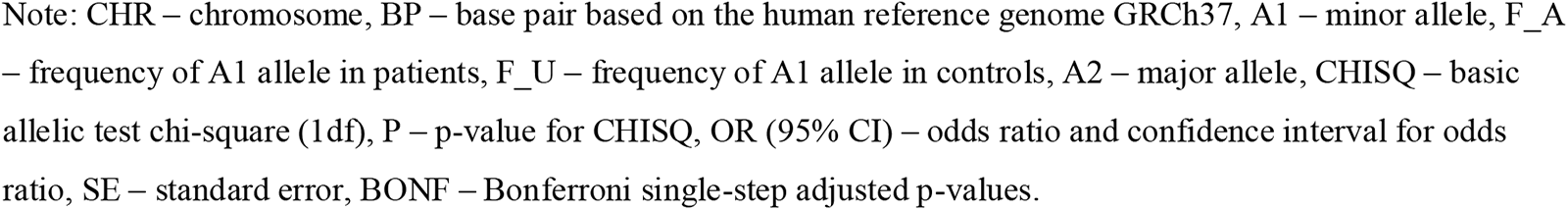
The exome-wide association study analysis of IgAN patients and control groups.

### Gene set enrichment and eQTL analysis

Using the Metascape tool, we built networks showing the specificities underlying the interactions between genes.

In additive model, the terms “CAMKK2 pathway” (WP4874) (log[q-value] = −0.27) and “positive regulation of catabolic process” (GO:0042176) (log[q-value] = −0.15) demonstrated the most significant overexpression (figure 3A).

In dominant model, the terms “cell-cell adhesion via plasma-membrane adhesion molecules” (GO:0098742) (log[q-value] = −0.19) and “ER-nucleus signaling pathway” (GO:0006984) (log[q-value] = −0.19) were significantly higher enriched (figure 3B).

In recessive model, the most overrepresented terms included “response to acid chemical” (GO:0001101) (log[q-value] = −0.97) and “MTOR signalling” (R-HSA-165159) (log[q-value] = −0.97) (figure 3C).

Moreover, cell type signature analysis revealed kidney-related terms to be the topmost. For instance, the “lake adult kidney c5 proximal tubule epithelial cells stress inflame” (M39224) was the first and the second hit for the recessive and additive models, respectively; for the recessive model, the second hit was “lake adult kidney c26 mesangial cells” (M39245). In case of the dominant model, the least significantly enriched terms were “lake adult kidney c11 thin ascending limb” (M39230) and “lake adult kidney c15 connecting tubule” (M39234).

For each inheritance model, we detected 4 variants: rs10710110 (*NBPF3*), rs34527214 (*CYTH2*), rs4803184 (*FBXO27*), and rs71185698 (*PSMD2*) (figure 3D). Based on the eQTL analysis relying on the NephQTL database, there was no substantial difference between the number of variants affecting the expression in the glomeruli and tubulointerstitium (figure 3E); however, their number was nearly two times higher than detected by DICE. We found only two variants in the Kidney eQTLs Atlas: rs2242954 affected *PWP2* expression in renal tubules and С*21orf33* expression in the renal tubules and glomeruli, while rs4803184 influenced *FBXO27* expression in the glomeruli. rs2242954 is also known to elevate the С*21orf33* expression in macrophages (beta = 18.259 ± 4.087, P = 1.156e^−5^), NK cells (beta = 9.086 ± 1.943, P = 4.593× 10^−6^), CD4+ Т cells (beta = 5.722 ± 1.312, P = 1.822 × 10^−5^), dendritic cells (beta = 4.964 ± 0.769, P = 4.818 × 10^−10^), and CD8+ Т cells (beta = 3.611 ± 0.689, P = 3.245 × 10^−7^).

**Figure 3.**
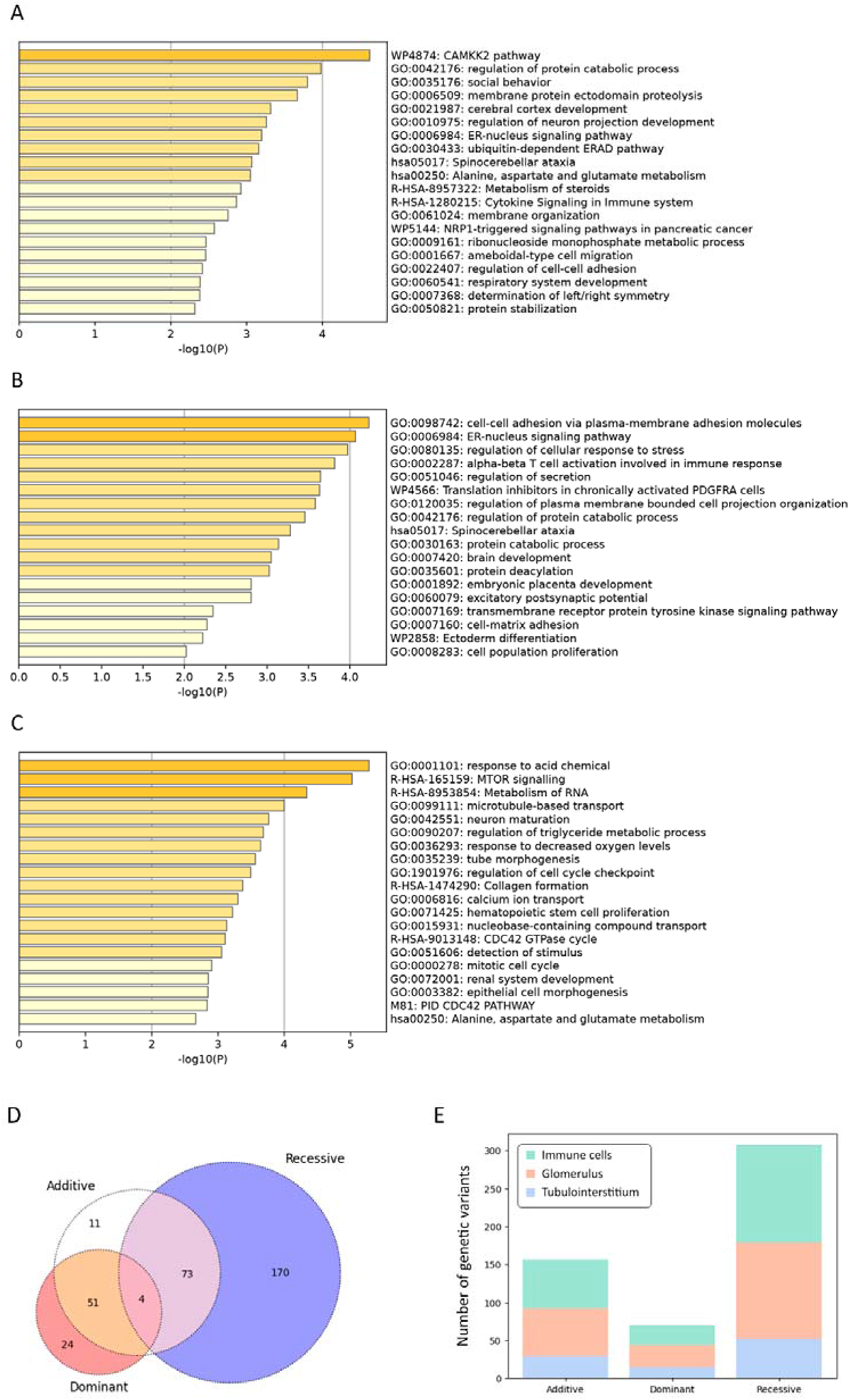
Statistically enriched terms using Metascape, additive (A), dominant (B) and recessive (C) logistic models. The intersection of genetic variant set based on different inheritance models, a Venn diagram (D). Markers are interrogated against the following datasets: Database of Immune Cell eQTLs (DICE) project and NephQTL (E).

### Protein-Protein Interaction (PPI) network analysis of potential targets

The PPI networks were obtained using the String online platform. None of the networks had PPI enrichment P < 0.05, the lowest value belonging to the recessive model with P = 0.0613. Its PPI contained 205 nodes, 160 edges, and the average node degree equal to 1.56. After clustering with k-means into 3 clusters, the most enriched cluster was the cluster of the recessive model with P = 3.09 × 10^−14^ (average node degree was 1.32) (figure 4).

**Figure 4.**
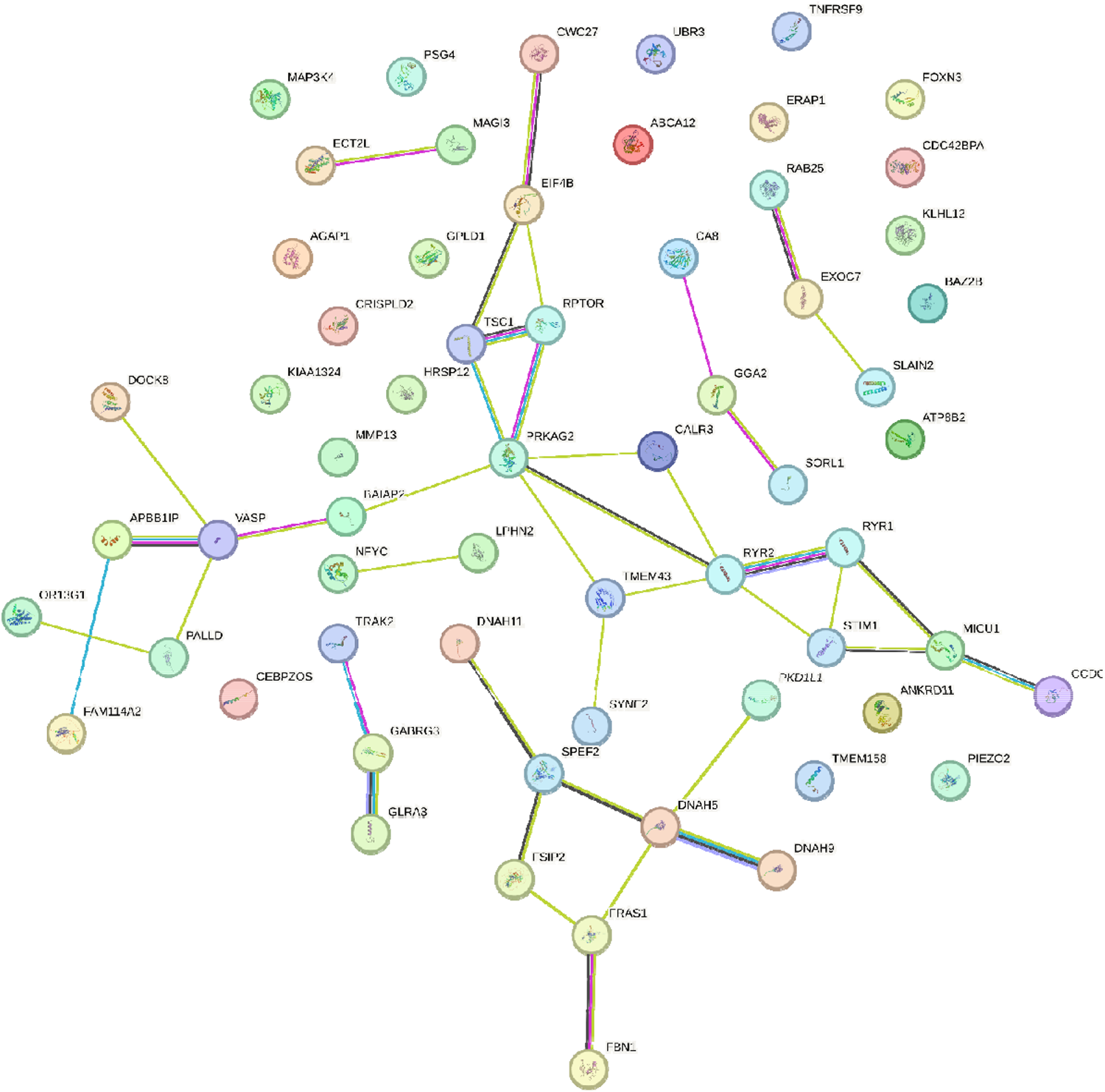
The STRING protein-protein interaction network based on gene interactions, recess ve model, one of the three clusters.

### Transcription factor (TF) binding sites’ enrichment analysis

We identified numerous regions with predicted TF binding sites near SNVs that could affect gene expression (table S3). 11 SNVs enhance and 2 SNVs disrupt transcription factor binding sites.

### Frequency and comparison of allele frequencies between IgAN patients and healthy donors

Table 3 shows the most frequent HLA alleles in the IgAN patient samples. Among them, we detected 142 unique alleles: 22 HLA-A, 34 HLA-B, 22 HLA-C, 28 HLA-DRB1, 7 HLA-DQA1, 13 HLA-DQB1, and 16 HLA-DPB1 alleles. The loci HLA-C (expected heterozygosit = 92.41%, observed heterozygosity = 90.00%, P = 0.0036) and HLA-DRB1 (expected heterozygosity = 92.65%, observed heterozygosity = 84.29%, P = 0.0013) exhibited statistically significant deviations from the Hardy-Weinberg equilibrium (table S4). In the healthy donors’ group, the most frequent alleles were HLA-A*02:01:01G (27.13%), HLA-B*07:02:01G (11.79%), HLA-C*07:02:01G (13.22%), HLA-DRB1*07:01:01G (13.36%), HLA-DQA1*05:01:01G (24.64%), HLA-DQB1*03:01:01G (22.12%) and HLA-DPB1*04:01:01G (44.19%) (table S5). Allele HLA-B*56:01:01G (OR, 4.331 [95% CI: 1.739-9.437]) demonstrated the strongest association with IgAN, although it did not reach statistical significance. In the Russian population, this allele was observed only among 1.5% of Nizhniy Novgorod citizens [62]. Alleles HLA-C*01:02:01G (P = 0.029; OR, 2.727 [95% CI, 1.477-4.738]), HLA-DRB1*01:01:01G (P = 0.034; OR, 2.189 [95% CI, 1.373-3.389]), HLA-DQA1*01:01:01G (P = 0.0076; OR, 2.021 [95% CI, 1.322-3.048]) and HLA-DQB1*05:01:01G (P = 0.01; OR, 2.124 [95% CI: 1.383-3.195]) were significantly more frequent among IgAN patients.

## Discussion

Our study is the first Russian EWAS conducted on children with IgAN that employs the same population-based control cohort, takes insertions/deletions into consideration as well as utilizes HLA typing with up to 3-field resolution. The limitation of the study is a relatively small patient cohort size, despite applying the strict Bonferroni criterion.

Epidemiological data on the IgAN frequency in Russia are scarce. According to the statistics from the period of 1999-2019, IgAN was the most common type of immune glomerulopathies (41.5%). It was detected in 1 out of every 4 kidney biopsies, indicating a higher prevalence compared to Asia, Europe, and America [63]. The annual incidence of IgAN is approximately 8-10 cases per 1 million, and the average age of the onset is 34±12 years. Therefore, the size of the children cohort prospectively recruited in Moscow for 3 years aligns with the expected size.

The most prominent signals were detected in the loci that had not been described before in IgAN patients. However, through eQTL analysis, we discovered that the variants affect gene expression in the glomeruli and renal tubules and confirmed their connection to significant Metascape categories related to immunity and kidney development. Moreover, the functional enrichment analysis revealed signaling pathways associated with nervous system development, which surprisingly had been already reported. *ANKRD16* was suggested as a candidate gene for IgAN in the Korean population [64], although its mutant protein was reported to be associated with the Purkinje cell degeneration [65] and had not been linked to the renal disorders before.

One of the examples that might help explain these findings is the *CYLD* gene. Its gain-of-function mutations was observed in the Alzheimer and Parkinson patients, while its loss-of-function mutations were discovered in the patients with the benign skin neoplasms, with no observed overlap between mutant phenotypes. Another possible explanation is phenotypic heterogeneity. We obtained the results comparable to the global data only by studying the HLA loci [17, 34]. In the patient group, we identified a slight predominance of the HLA-DRB1∗03:01:01 [26] and HLA-DQB1*03:01 [28] alleles, although it was not statistically significant.

The rs143409664 variant is VUS since it produces an elongated protein while the alteration in the nucleotide sequence does not reside in the regions with repeats. In the Russian population, the rs143409664-C allele has frequencies of 0.5211 among healthy individuals and 0.5242 among the patients according to RuSeq [67]. *PRAG1* knockout in mice is lethal [68], and this gene has been described in several papers. Although its expression was detected in kidneys as well [69], there is no reliable evidence supporting the association of *PRAG1* with the disease.

The frequency of the rs5828672 variant in the *ZNF787* gene is 0.9085, however, it was absent in RuSeq, which may indirectly imply the inexplicable predominance of the allele in IgAN patients. This variant did not show any linkage disequilibrium as well. *ZNF787* has been characterized to a certain extent; it’ possesses a proven ability (it is known to be able) to suppress the neuron growth and differentiation from iPSC [70]. The frequency of the *UBR3* gene rs13028230-G allele is 0.639, according to gnomAD, indicating the lower representation of the A allele; however, no information on it could be found in RuSeq. The highest value of r^2^, the indicator of the linkage disequilibrium, in our sample belonged to rs72891954 (0.327), which slightly deviates from the value in the global population (0.282, P < 0.0001) [71]. Therefore, the hypothesis suggesting that the UBR3 allele belongs to a haplotype due to population shifts does not appear convincing. The UBR3 mRNA levels in patients with lupus nephritis correlated with SLEDAI-2K and the index of histological activity [72]. The repertoires of the B and T cell receptors were studied in IgAN patients, and some types were found to be associated with the disease [73]. We identified the previously undescribed variant rs767448033 in the *TRBV5-4* gene. The expression of the transcription factor EOMES was higher in the circulating CD8 T cells (59%) compared to CD4 T cells (15%); however, in the kidney tissue, its expression in CD4 T cells was higher than in blood (32%) [74]. An elevated SREBF1 expression was detected in patients with the chronic renal disease [75]. The *CHRNA3* gene variants correlated with eGFR, albumin-creatinine ratio, and albuminuria. Although *OR10G6* gene is related to the olfactory system, its variant rs1453654 was associated with the elevated expression of IFNγ after smallpox vaccination [77]. In the rat model of hypertension disease, Piezo2 activation was observed in mesangial cells, renin cells, and perivascular mesenchymal cells implying its contribution to renal fibrosis [78]. Based on the dominant model, we identified the *ITGAM* gene among the IgAN candidate genes with repeatedly confirmed association [79]. Its variant rs60662530 had P_BONF_ = 0.046, however, the frequency ratio did not support the suggestion that the -CTTG allele was more frequent among patients (OR, 2.887 [95% CI: 0.1893-1.992]), as the lower CI was less than 1.

We did not study the external risk factors, therefore, we can rely only on the ratio of the allele frequencies and genotypes from the samples.

Although there are quite a few studies on genetics of IgAN patients, the role of particular genes as well as signaling pathways in the disease pathogenesis remains elusive. This complexity may be attributed to the highly heterogeneous interactions of hereditary factors or ethnospecificity of their manifestation. Answering these questions require further research on larger samples utilizing more powerful molecular and genetic techniques such as GWAS and transcriptome comparison.

## Data availability statement

The data underlying this article will be shared on reasonable request to the corresponding author.

## Authors’ contributions

Conceptualization: MVP, EKP; formal analysis: AAB, VVC; visualization: AAB; resources: MVP, DOK; software: VVC, ASP, IAV, ONS; acquisition of data: VAB, AOS; project administration: EKP, DOK; supervision: DVR; writing – original draft preparation: AAB, MVP; writing – review & editing: EKP, DOK. Each author contributed important intellectual content during manuscript drafting or revision and agrees to be personally accountable for the individual’s own contributions and to ensure that questions pertaining to the accuracy or integrity of any portion of the work, even one in which the author was not directly involved, are appropriately investigated and resolved, including with documentation in the literature if appropriate.

## Supplementary Material

**Supplementary Figure 1A.**
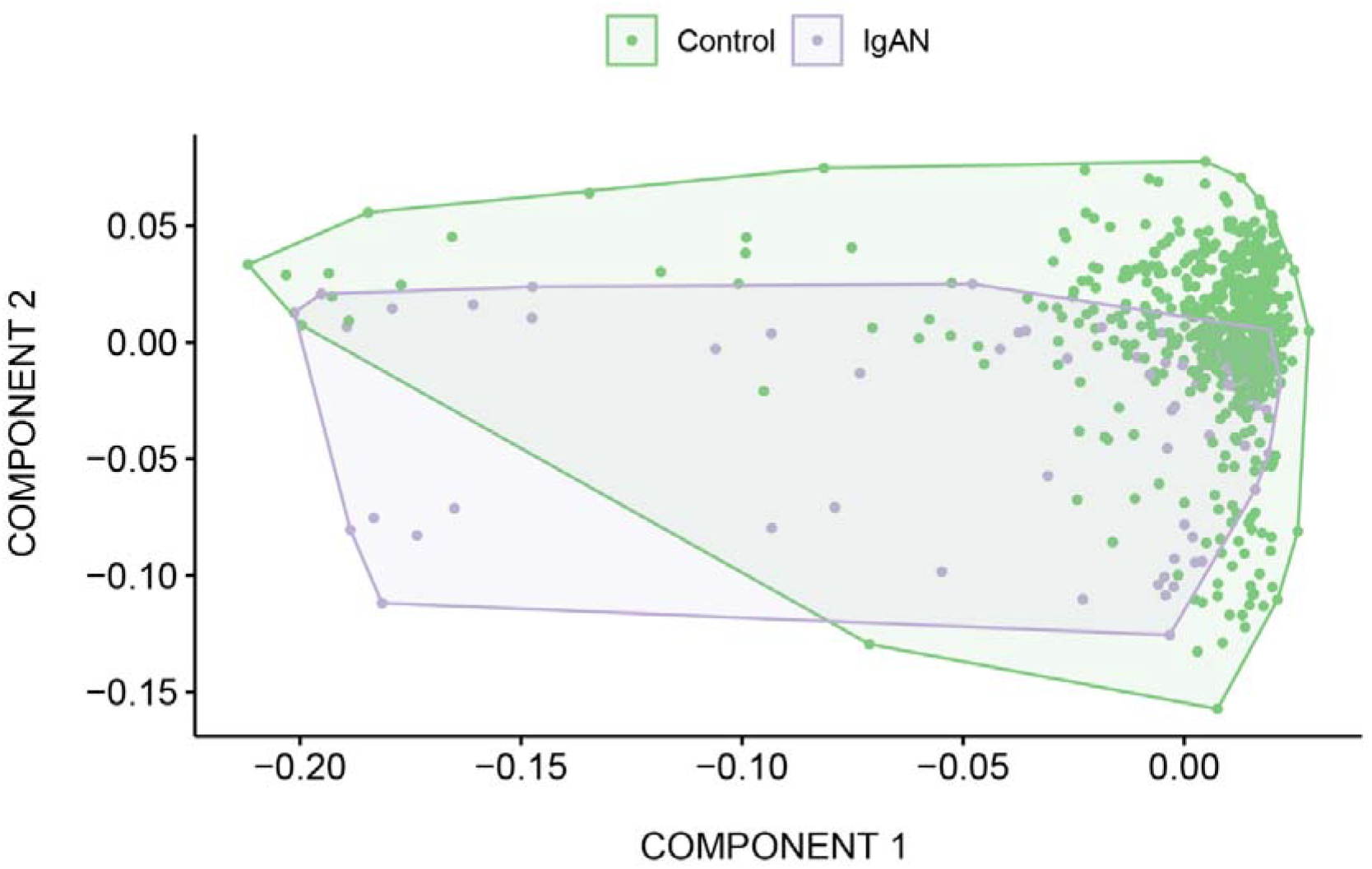
Principal component analysis of the IgAN samples comprising the samples from healthy donor cohort.

**Supplementary Figure 1B.**
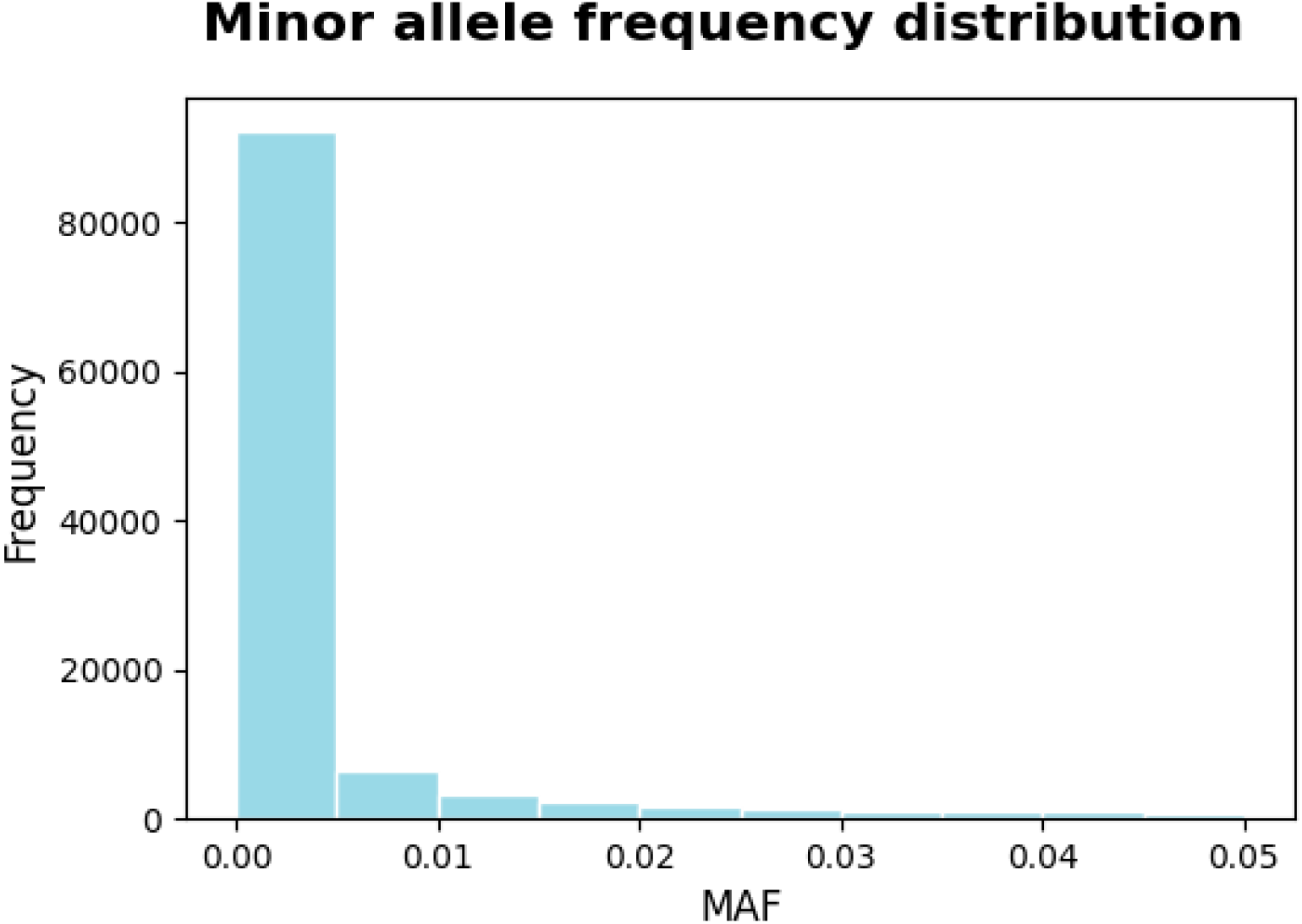
Minor allele frequency distribution in IgAN patients and healthy donors.

**Supplementary Table 1.**
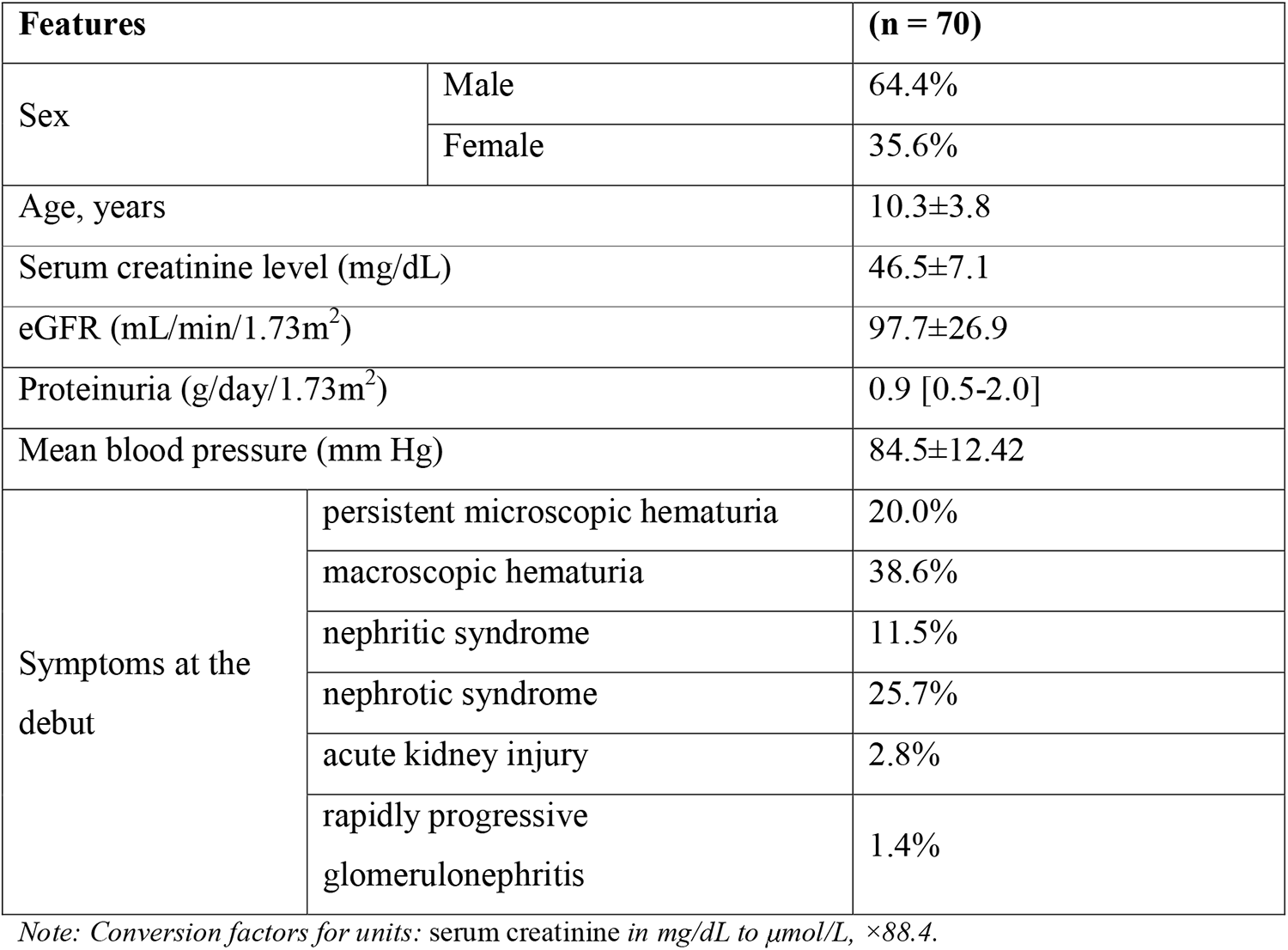
Clinical characteristics of the IgAN patients.

**Supplementary Table 2.** Key quality control metrics for exome sequencing of the experimental samples from the IgAN cohort.

| Metrics | Mean | Min | Max |
| --- | --- | --- | --- |
| Single reads per sample | 79,494,574 | 41,389,840 | 162,054,108 |
| Estimated library size | 167,316,756 | 43,904,883 | 362,437,565 |
| Duplicates | 12.33 | 5.86 | 26.82 |
| On-target bases | 85.3% | 62% | 92% |
| Mean target coverage | 89.65 | 46.2 | 140 |
| Median target coverage | 80.62 | 42 | 128 |
| Width 10X | 96.9% | 94.3% | 97% |
| Width 20X | 95.2% | 85% | 97% |
| Width 30X | 91.2% | 70% | 96% |

**Supplementary Table 3.**
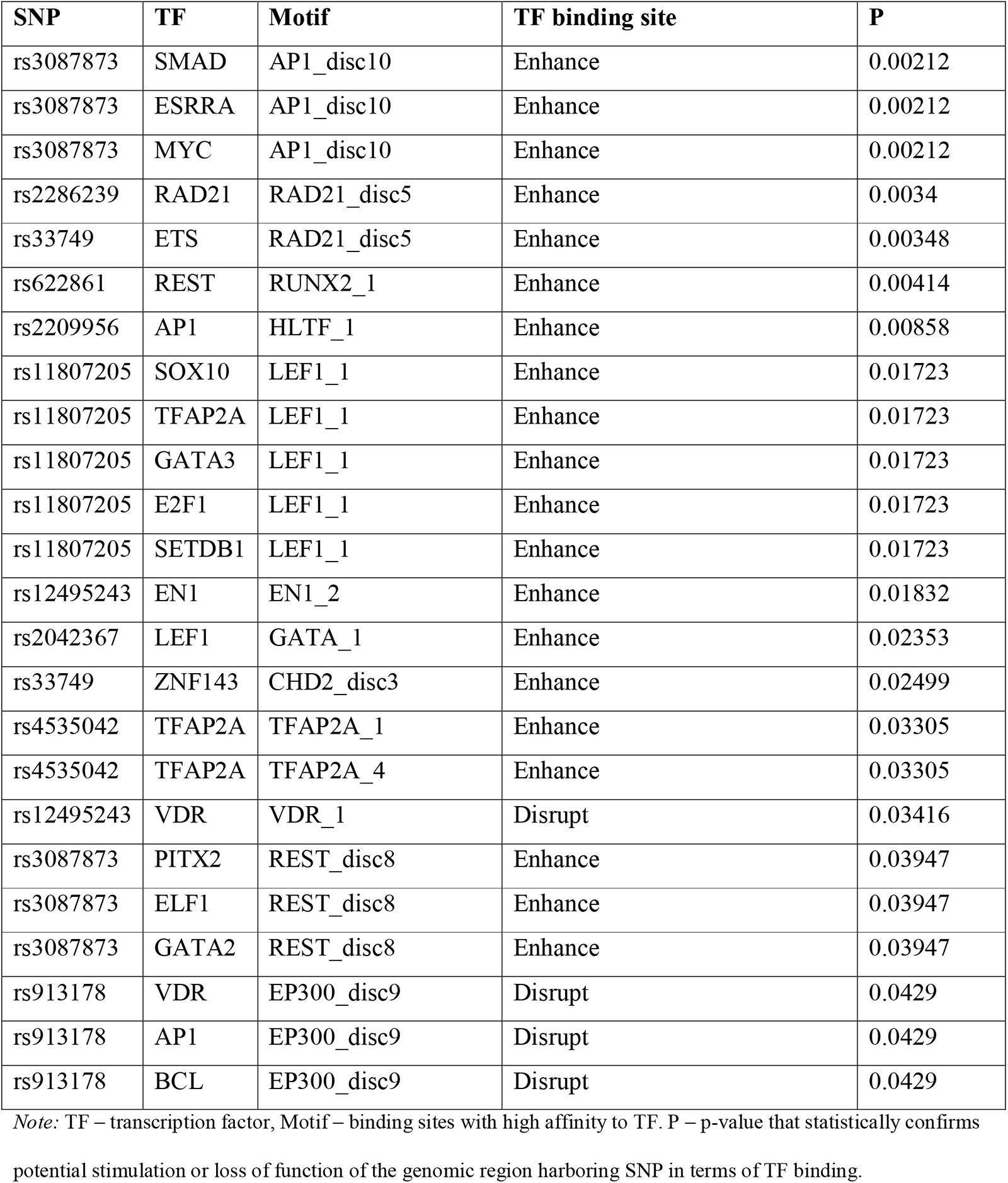
The TF binding motifs affected by potentially deleterious variants.

**Supplementary Table 4.** Conformity to Hardy-Weinberg equilibrium.

| Locus | No. of Genotypes | Observed Heterozygosity | Expected Heterozygosity | p-value |
| --- | --- | --- | --- | --- |
| IgAN |  |  |  |  |
| *HLA-A* | 70 | 0.88571 | 0.86228 | 0.41045 |
| *HLA-B* | 70 | 0.92857 | 0.96310 | 0.18927 |
| *HLA-C* | 70 | 0.90000 | 0.92405 | 0.00358 |
| *HLA-DRB1* | 70 | 0.84286 | 0.92652 | 0.00125 |
| *HLA-DQA1* | 70 | 0.67143 | 0.78335 | 0.05311 |
| *HLA-DQA2* | 70 | 0.72857 | 0.83659 | 0.09385 |
| *HLA-DPB1* | 70 | 0.78571 | 0.81408 | 0.18547 |

**Supplementary Table 5.**
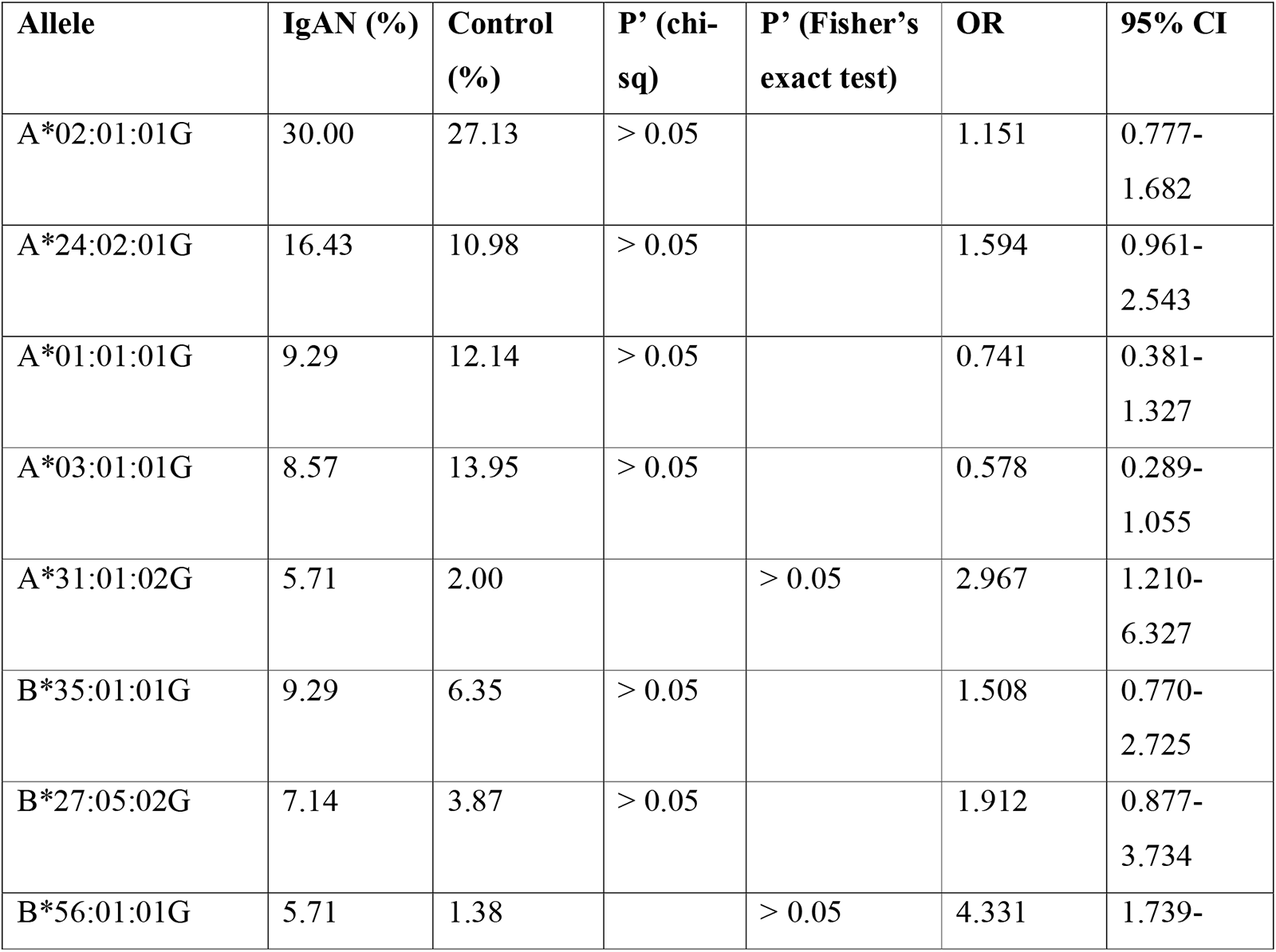
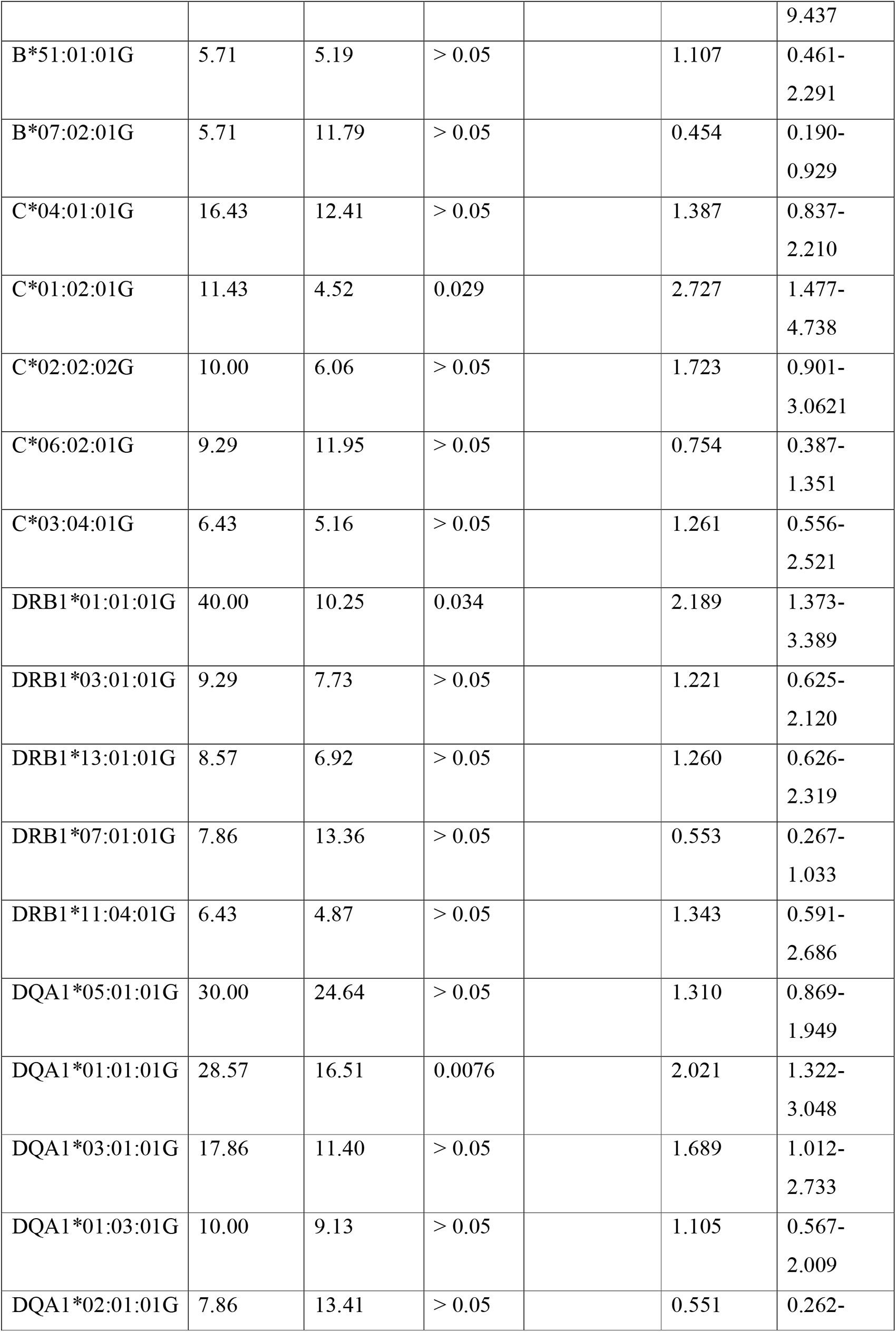
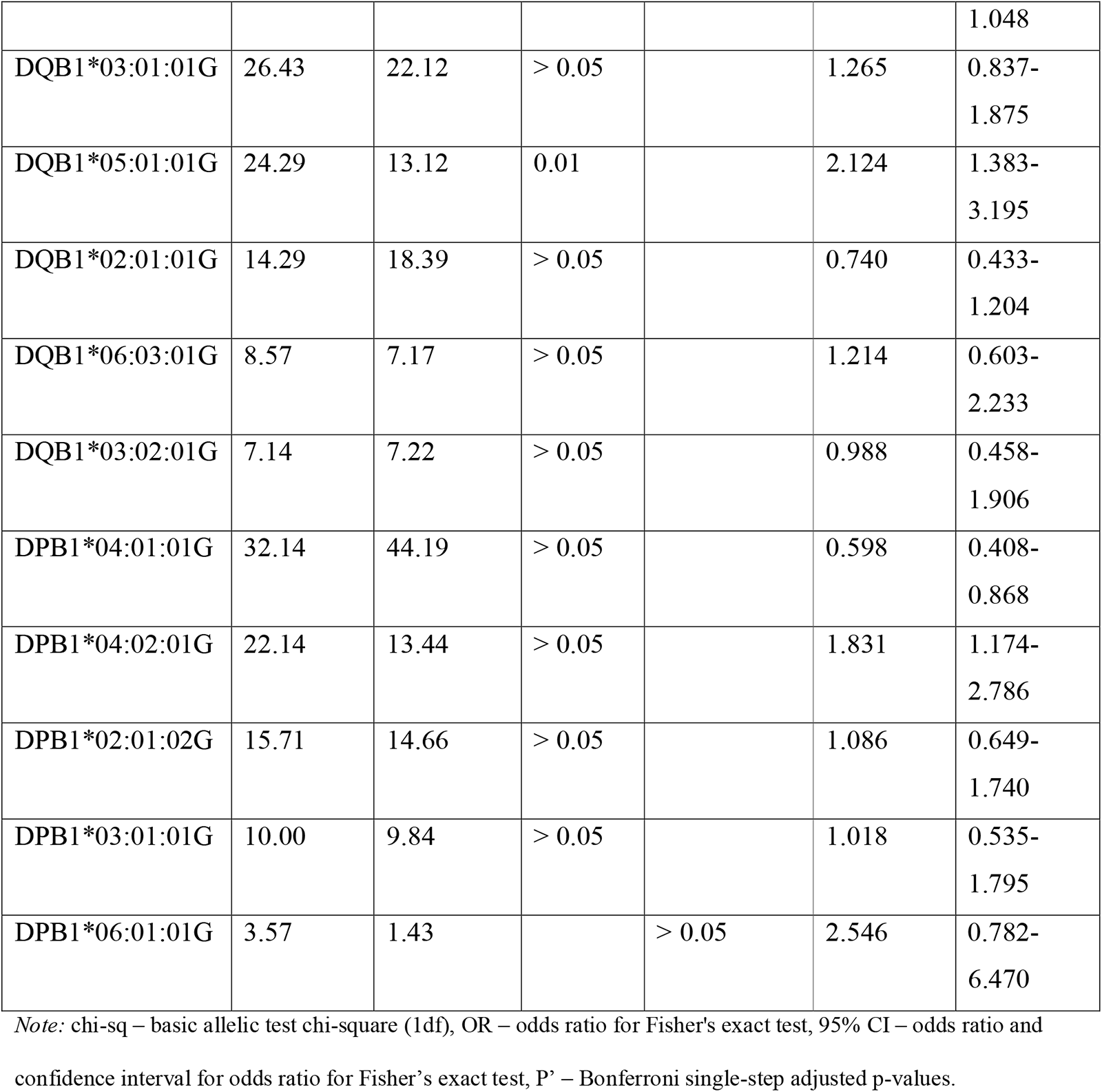
The distribution of the alleles HLA-A, -B, -DRB1, -DQA1, -DQB1 and -DPB1 in IgAN patients and healthy donors.

